# Effect of BNT162b2 antigen dosage on protection against SARS-CoV-2 omicron infection

**DOI:** 10.1101/2022.11.29.22282864

**Authors:** Hiam Chemaitelly, Houssein H. Ayoub, Peter Coyle, Patrick Tang, Hadi M. Yassine, Asmaa A. Al Thani, Hebah A. Al-Khatib, Mohammad R. Hasan, Zaina Al-Kanaani, Einas Al-Kuwari, Andrew Jeremijenko, Anvar Hassan Kaleeckal, Ali Nizar Latif, Riyazuddin Mohammad Shaik, Hanan F. Abdul-Rahim, Gheyath K. Nasrallah, Mohamed Ghaith Al-Kuwari, Hamad Eid Al-Romaihi, Adeel A. Butt, Mohamed H. Al-Thani, Abdullatif Al-Khal, Roberto Bertollini, Laith J. Abu-Raddad

**Affiliations:** Infectious Disease Epidemiology Group, Weill Cornell Medicine-Qatar, Cornell University, Doha, Qatar; World Health Organization Collaborating Center for Disease Epidemiology Analytics on HIV/AIDS, Sexually Transmitted Infections, and Viral Hepatitis, Weill Cornell Medicine–Qatar, Cornell University, Qatar Foundation – Education City, Doha, Qatar; Department of Population Health Sciences, Weill Cornell Medicine, Cornell University, New York, New York, USA; Mathematics Program, Department of Mathematics, Statistics, and Physics, College of Arts and Sciences, Qatar University, Doha, Qatar; Hamad Medical Corporation, Doha, Qatar; Biomedical Research Center, QU Health, Qatar University, Doha, Qatar; Wellcome-Wolfson Institute for Experimental Medicine, Queens University, Belfast, United Kingdom; Department of Pathology, Sidra Medicine, Doha, Qatar; Department of Biomedical Science, College of Health Sciences, QU Health, Qatar University, Doha, Qatar; Department of Public Health, College of Health Sciences, QU Health, Qatar University, Doha, Qatar; Primary Health Care Corporation, Doha, Qatar; Ministry of Public Health, Doha, Qatar; Department of Medicine, Weill Cornell Medicine, Cornell University, New York, New York, USA

**Keywords:** COVID-19, antibody, variant, omicron, vaccination, cohort study, immunity, epidemiology

## Abstract

**Background:** Coronavirus Disease 2019 (COVID-19) vaccine antigen dosage may affect protection against severe acute respiratory syndrome coronavirus 2 (SARS-CoV-2) infection, but direct evidence to quantify this effect is lacking.

**Methods:** A matched, retrospective, cohort study that emulated a randomized control trial was conducted in Qatar between February 3, 2022 and November 8, 2022, to provide a head-to-head, controlled comparison of protection induced by two antigen dosages of the BNT162b2 vaccine. The study compared incidence of omicron infection in the national cohort of adolescents 12 years of age who received the two-dose primary-series of the 30-µg BNT162b2 vaccine to that in the national cohort of adolescents 11 years of age who received the two-dose primary-series of the pediatric 10-µg BNT162b2 vaccine. Associations were estimated using Cox proportional-hazard regression models.

**Results:** Among adolescents with no record of prior infection, cumulative incidence of infection was 6.0% (95% CI: 4.9-7.3%) for the 30-µg cohort and 7.2% (95% CI: 6.1-8.5%) for the 10-µg cohort, 210 days after the start of follow-up. Incidence during follow-up was dominated by omicron subvariants including, consecutively, BA.1/BA.2, BA.4/BA.5, BA.2.75*, and XBB. The adjusted hazard ratio comparing incidence of infection in the 30-µg cohort to the 10-µg cohort was 0.77 (95% CI: 0.60-0.98). Corresponding relative effectiveness was 23.4% (95% CI: 1.6-40.4%). Relative effectiveness was -3.3% (95% CI: -68.0-27.5%) among adolescents with a record of prior infection.

**Conclusions:** Three-fold higher BNT162b2 dosage was associated with ∼25% higher protection against infection in infection-naïve adolescents of similar age. These findings may inform design of future COVID-19 vaccines and boosters for persons of different age groups.

## Introduction

The effect of antigen dosage of mRNA Coronavirus Disease 2019 (COVID-19) vaccines on protection against infection with severe acute respiratory syndrome coronavirus 2 (SARS-CoV-2) is poorly understood. The observed differences in the protection of the mRNA-1273^1^ (Moderna) vaccine and the BNT162b2^2^ (Pfizer-BioNTech) vaccine suggest a role for antigen dosage in determining vaccine protection.^3^ The observed differences in vaccine effectiveness among children vaccinated with the pediatric 10-µg BNT162b2 dosage compared to adolescents vaccinated with the 30-µg BNT162b2 dosage suggests also an effect for antigen dosage in determining vaccine protection.^4^ However, no studies to date have directly investigated the effect of antigen dosage on protection using a head-to-head controlled comparison of the protection induced by two different antigen dosages of the same vaccine.

We conducted a matched, retrospective, target-trial, cohort study in Qatar that compares head-to-head the relative effectiveness against SARS-CoV-2 omicron (B.1.1.529) infection of the primary-series of the 30-µg antigen dosage among adolescents 12 years of age to that of the 10-µg antigen dosage among adolescents 11 years of age. This design fundamentally emulates a retrospective, unblinded, controlled trial.^5,6^ While the study should have ideally been implemented among persons of the same exact age, the age difference of one year between the study cohorts should have a minimal effect on study findings, and in the direction of the null hypothesis of no difference in protection, since vaccine effectiveness of the same antigen dosage appears stronger at younger age.^4^ Following the US Food and Drug Administration guidelines, the 30-µg BNT162b2 antigen dosage was authorized in Qatar for persons ≥12 years of age while the 10-µg BNT162b2 antigen dosage was authorized for children 5-11 years of age.^2,7^

## Methods

### Study population and data sources

This study was conducted between February 3, 2022, the earliest record for vaccination with the pediatric 10-µg BNT162b2 vaccine, and November 8, 2022. It analyzed the national, federated databases for COVID-19 laboratory testing, vaccination, hospitalization, and death, retrieved from the integrated, nationwide, digital-health information platform (Section S1 of the Supplementary Appendix). Databases include all SARS-CoV-2-related data with no missing information since the onset of the pandemic, including all polymerase chain reaction (PCR) tests, and from January 5, 2022 onward, all rapid antigen tests conducted at healthcare facilities (Section S2). SARS-CoV-2 testing in Qatar has been done at mass scale up to October 31, 2022, mostly for routine reasons.^8,9^ Most infections were diagnosed not because of symptoms, but because of routine testing.^8,9^ Qatar launched its mass COVID-19 immunization campaigns in adolescents ≥12 years of age in February of 2021 using the 30-µg BNT162b2 antigen dosage, and in children 5-11 years of age in February of 2022 using the pediatric 10-µg BNT162b2 antigen dosage.^4^ Detailed descriptions of Qatar’s population and of the national databases have been reported previously.^4,6,8-11^

### Study design and cohorts

A matched, retrospective, cohort study that emulated a randomized control trial^5,6^ was conducted to estimate the relative effectiveness against SARS-CoV-2 omicron infection of the primary-series of the 30-µg antigen dosage among adolescents 12 years of age compared to that of the 10-µg antigen dosage among adolescents 11 years of age.

The study compared incidence of omicron infection in the national cohort of adolescents 12 years of age who received the two-dose primary-series of the 30-µg BNT162b2 vaccine (designated the 30-µg cohort) to that in the national control cohort of adolescents 11 years of age who received the two-dose primary-series of the pediatric 10-µg BNT162b2 vaccine (designated the 10-µg cohort).

Documentation of infection in all cohorts was based on positive PCR or rapid antigen tests, regardless of symptoms. Laboratory methods are found in Section S2. Classification of COVID-19 infection severity (acute-care hospitalizations),^12^ criticality (intensive-care-unit hospitalizations),^12^ and fatality^13^ followed World Health Organization guidelines (Section S3).

### Cohort matching and follow-up

Any 12-year-old adolescent was eligible for inclusion in the 30-µg cohort if the adolescent received two doses of the 30-µg BNT162b2 vaccine, completed 14 days after the second dose, and had no record for a SARS-CoV-2-positive test within 90 days of the start date of follow-up. The latter exclusion criterion ensured that infections after start of follow-up were incident infections and not prolonged SARS-CoV-2-positivity of earlier infections.^14-16^

Adolescents in the 30-µg cohort were matched exactly one-to-one by sex, 10 nationality groups, number of coexisting conditions (none or one, two or more), prior infection status (no prior infection, or prior infection with either pre-omicron or omicron viruses, or prior infections with both viruses) to adolescents in the 10-µg cohort, to balance observed confounders between exposure groups related to infection risk in Qatar.^10,17-20^ Matching by these factors was shown previously to provide adequate control of differences in SARS-CoV-2 infection risk.^3,9,21-23^

Matching was also done by calendar month of the second vaccine dose. That is, matched pairs needed to have received their second dose during the same calendar month. This matching was done to control for time since second-dose vaccination and to ensure that each matched pair was exposed to the same force of infection and omicron subvariants and had a record of active presence in Qatar during the same calendar time. Matching was performed iteratively so that adolescents in the 10-µg cohort were alive and had no record before the start date of follow-up for a SARS-CoV-2-positive test within 90 days. Iterations were performed until no more matched pairs could be identified.

Each matched pair was followed from the calendar day when the adolescent in the 30-µg cohort completed 14 days after the second dose. To ensure exchangeability,^6,24^ both members of each matched pair were censored on the earliest occurrence of an adolescent in either cohort receiving a third (booster) vaccine dose. Accordingly, adolescents were followed up until the first of any of the following events: a documented SARS-CoV-2 infection, or third-dose vaccination (with matched pair censoring), or death, or end of study censoring (November 8, 2022).

### Oversight

The institutional review boards at Hamad Medical Corporation and Weill Cornell Medicine– Qatar approved this retrospective study with a waiver of informed consent. The study was reported according to the Strengthening the Reporting of Observational Studies in Epidemiology (STROBE) guidelines (Table S1). The authors vouch for the accuracy and completeness of the data and for the fidelity of the study to the protocol. Data used in this study are the property of the Ministry of Public Health of Qatar and were provided to the researchers through a restricted-access agreement for preservation of confidentiality of patient data. The funders had no role in the study design; the collection, analysis, or interpretation of the data; or the writing of the manuscript.

### Statistical analysis

Eligible and matched cohorts were described using frequency distributions and measures of central tendency and were compared using standardized mean differences (SMDs). An SMD of ≤0.1 indicated adequate matching.^25^ Cumulative incidence of infection (defined as the proportion of individuals at risk, whose primary endpoint during follow-up was an infection) was estimated using the Kaplan–Meier estimator method.^26^ Incidence rate of infection in each cohort, defined as the number of identified infections divided by the number of person-weeks contributed by all individuals in the cohort, was estimated, along with its 95% confidence interval (CI), using a Poisson log-likelihood regression model with the Stata 17.0 *stptime* command.

The hazard ratio (HR), comparing incidence of infection in the cohorts and corresponding 95% CIs, was calculated using Cox regression adjusted for the matching factors with the Stata 17.0 *stcox* command. Sensitivity analysis further adjusting the HR for differences in testing frequency between cohorts was conducted. Schoenfeld residuals and log-log plots for survival curves were used to test the proportional-hazards assumption and to investigate its adequacy. 95% CIs were not adjusted for multiplicity; thus, they should not be used to infer definitive differences between groups. Interactions were not considered.

Relative vaccine effectiveness was estimated as 1-adjusted hazard ratio (aHR) if the aHR was <1, and as 1/aHR-1 if the aHR was ≥1. These expressions were used to ensure symmetric scale for both negative and positive relative effectiveness, ranging from -100%-100%.

The main analysis investigated the relative effectiveness by restricting the cohorts to persons with no record of prior infection before the start of the follow-up. This was done to disentangle the protection induced by only vaccination from that induced by hybrid immunity of vaccination and natural infection.^8,27^

Additional analyses were conducted to investigate the relative effectiveness in adolescents with prior pre-omicron or omicron infections and in the full matched cohorts of adolescents regardless of prior infection status. Statistical analyses were conducted using Stata/SE version 17.0 (Stata Corporation, College Station, TX, USA).

## Results

### Study population

Figure S1 shows the study population selection process. Table 1 describes the baseline characteristics of the full study population and matched cohorts. Matched cohorts each included 2,999 persons.

**Table 1.** Baseline characteristics of the eligible and matched cohorts in the study investigating relative effectiveness of the 30-µg BNT162b2 vaccine compared to the pediatric 10-µg BNT162b2 vaccine against infection with an omicron subvariant.

| Characteristics | Eligible cohorts |  |  | Matched cohorts* |  |  | Matched cohorts* |  |  |
| --- | --- | --- | --- | --- | --- | --- | --- | --- | --- |
|  |  |  |  | No prior infection |  |  | Prior infection |  |  |
| | 30-μg cohort | 10-μg cohort | SMD† | 30-μg cohort | 10-μg cohort | SMD† | 30-μg cohort | 10-μg cohort | SMD† |
|  | N=4,085 | N=5,323 |  | N=2,460 | N=2,460 |  | N=539 | N=539 |  |
| Sex |  |  |  |  |  |  |  |  |  |
| Male | 2,058 (50.4) | 2,771 (52.1) | 0.03 | 1,255 (51.0) | 1,255 (51.0) | 0.00 | 275 (51.0) | 275 (51.0) | 0.00 |
| Female | 2,027 (49.6) | 2,552 (47.9) |  | 1,205 (49.0) | 1,205 (49.0) |  | 264 (49.0) | 264 (49.0) |  |
| Nationality‡ |  |  |  |  |  |  |  |  |  |
| Bangladeshi | 79 (1.9) | 161 (3.0) | 0.47 | 50 (2.0) | 50 (2.0) | 0.00 | 7 (1.3) | 7 (1.3) | 0.00 |
| Egyptian | 652 (16.0) | 396 (7.4) |  | 295 (12.0) | 295 (12.0) |  | 57 (10.6) | 57 (10.6) |  |
| Filipino | 281 (6.9) | 584 (11.0) |  | 183 (7.4) | 183 (7.4) |  | 61 (11.3) | 61 (11.3) |  |
| Indian | 1,469 (36.0) | 2,664 (50.0) |  | 1,082 (44.0) | 1,082 (44.0) |  | 269 (49.9) | 269 (49.9) |  |
| Nepalese | 10 (0.2) | 19 (0.4) |  | 3 (0.1) | 3 (0.1) |  | -- | -- |  |
| Pakistani | 306 (7.5) | 376 (7.1) |  | 188 (7.6) | 188 (7.6) |  | 32 (5.9) | 32 (5.9) |  |
| Qatari | 206 (5.0) | 86 (1.6) |  | 60 (2.4) | 60 (2.4) |  | -- | -- |  |
| Sri Lankan | 61 (1.5) | 105 (2.0) |  | 37 (1.5) | 37 (1.5) |  | 12 (2.2) | 12 (2.2) |  |
| Sudanese | 118 (2.9) | 43 (0.8) |  | 31 (1.3) | 31 (1.3) |  | 1 (0.2) | 1 (0.2) |  |
| Other nationalities§ | 903 (22.1) | 889 (16.7) |  | 531 (21.6) | 531 (21.6) |  | 100 (18.6) | 100 (18.6) |  |
| Number of coexisting conditions |  |  |  |  |  |  |  |  |  |
| 0-1 | 3,968 (97.1) | 5,192 (97.5) | 0.03 | 2,425 (98.6) | 2,425 (98.6) | 0.00 | 536 (99.4) | 536 (99.4) | 0.00 |
| 2+ | 117 (2.9) | 131 (2.5) |  | 35 (1.4) | 35 (1.4) |  | 3 (0.6) | 3 (0.6) |  |
| Prior infection |  |  |  |  |  |  |  |  |  |
| No prior infection | 3,207 (78.5) | --¶ | -- | 2,460 (100.0) | 2,460 (100.0) | -- | -- | -- | 0.00 |
| Pre-Omicron prior infection | 503 (12.3) | --¶ |  | -- | -- |  | 289 (53.6) | 289 (53.6) |  |
| Omicron prior infection | 360 (8.8) | --¶ |  | -- | -- |  | 246 (45.6) | 246 (45.6) |  |
| Pre-Omicron & Omicron prior infections | 15 (0.4) | --¶ |  | -- | -- |  | 4 (0.7) | 4 (0.7) |  |

IQR denotes interquartile range, SARS-CoV-2 severe acute respiratory syndrome coronavirus 2, and SMD standardized mean difference.
\*Each adolescent vaccinated with the 30-μg BNT162b2 vaccine was matched exactly one-to-one by sex, 10 nationality groups, number of coexisting conditions, prior infection status, and calendar month of the second vaccine dose to the first eligible adolescent vaccinated with the pediatric 10-μg BNT162b2 vaccine who was alive and did not have a SARS-CoV-2 positive test in the 90 days before the start of the follow-up (14 days after the second vaccine dose of their match).
†SMD is the difference in the mean of a covariate between groups divided by the pooled standard deviation. An SMD $\leq 0.1$ indicates adequate matching.
‡Nationalities were chosen to represent the most populous groups in Qatar.
§These comprise up to 74 other nationalities in the unmatched cohorts, and up to 64 other nationalities in the matched cohorts with no prior infection and up to 33 other nationalities in the matched cohorts with prior infection.
¶Ascertained at the start of follow-up. Accordingly, distribution is not available for the unmatched 10-μg cohort, as the start of follow-up for each person in the 10-μg cohort is determined by that of their match in the 30-μg cohort (14 days after the second dose) after the matching is performed.

### Relative effectiveness in adolescents with no prior infection

The matched cohorts restricted to persons with no prior infection included each 2,460 adolescents (Figure S1). For the 30-µg cohort, the median date of the first dose was March 29, 2022 and of the second dose was April 27, 2022. The median duration between the first and second doses was 28 days (interquartile range (IQR), 21-28 days).

For the 10-µg cohort, the median date of the first dose was March 31, 2022 and of the second dose was April 22, 2022. The median duration between the first and second doses was 21 days (IQR, 21-21 days). The median difference in age between the matched adolescents in the 30-µg and 10-µg cohorts was 8.4 months (IQR, 5.2-11.6 months).

The median duration of follow-up was 181 days (IQR, 137-216 days) for the 30-µg cohort and 179 days (IQR, 136-216 days) for the 10-µg cohort (Figure 1). During follow-up, 109 infections were recorded in the 30-µg cohort and 140 infections were recorded in the 10-µg cohort (Figure S1 and Table 2). None of the infections progressed to severe,^12^ critical,^12^ and fatal^13^ COVID-19.

**Figure 1.**
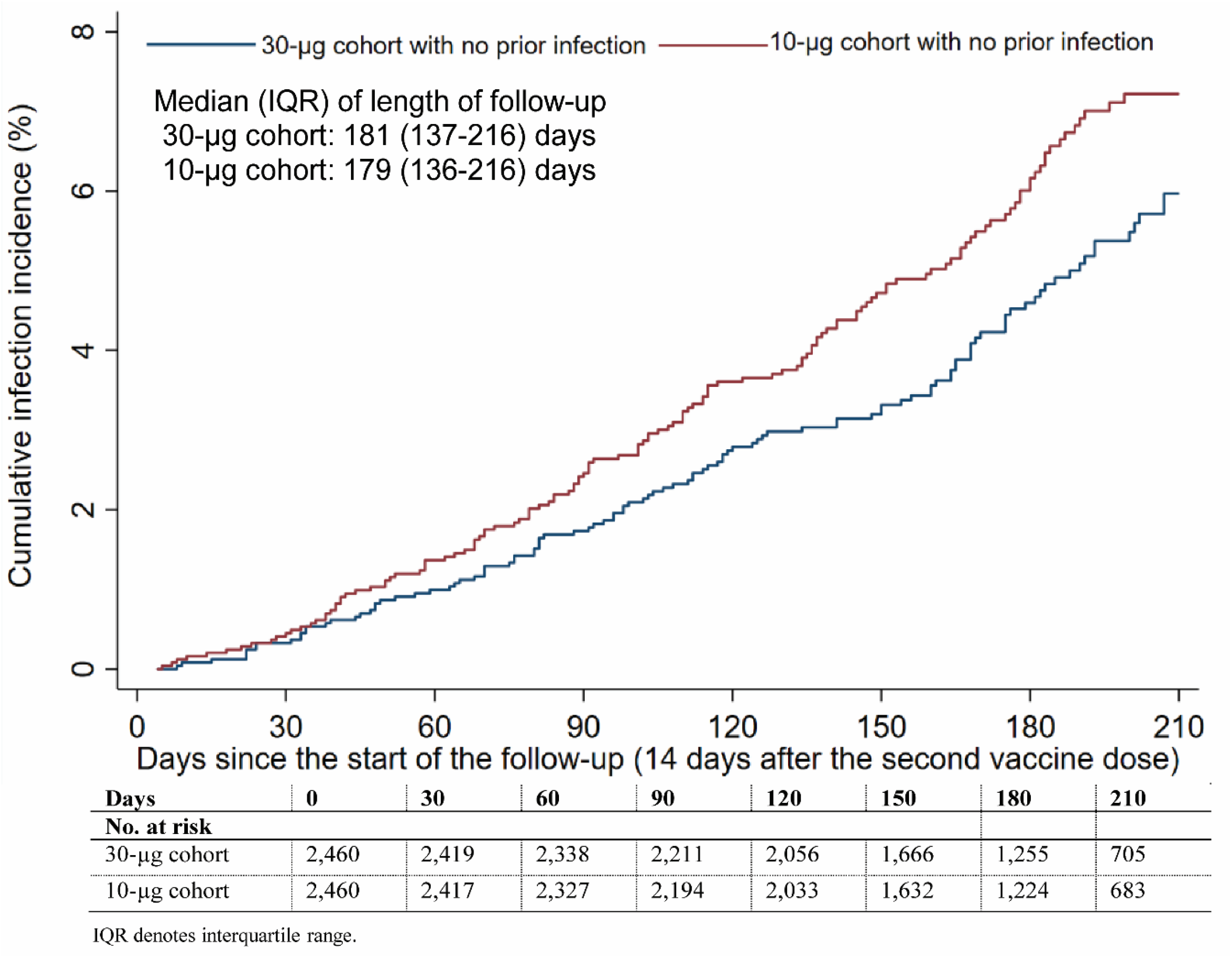
Cumulative incidence of SARS-CoV-2 omicron infection in adolescents who received two doses of the 30-µg BNT162b2 vaccine compared to those who received two doses of the pediatric 10-µg BNT162b2 vaccine.

**Table 2.** Adjusted hazard ratio and relative vaccine effectiveness against omicron infection in the 30-µg versus the 10-µg cohorts.

| **Epidemiological measures** | **30-µg cohort\*** | **10-µg cohort\*** |
| --- | --- | --- |
| **Main analysis-Matched cohorts with no prior infection** |  |  |
| Total follow-up time (person-weeks) | 60,230 | 59,604 |
| Number of incident infections | 109 | 140 |
| Incidence rate of infection (per 10,000 person-weeks; 95% CI) | 18.1 (15.0 to 21.8) | 23.5 (19.9 to 27.7) |
| Unadjusted hazard ratio for infection with an omicron subvariant (95% CI) | 0.77 (0.60 to 0.99) |  |
| Adjusted hazard ratio for infection with an omicron subvariant (95% CI)<sup>†</sup> | 0.77 (0.60 to 0.98) |  |
| Effectiveness against infection with an omicron subvariant in % (95% CI)<sup>‡</sup> | 23.4 (1.6 to 40.4) |  |
| **Additional analyses** |  |  |
| ***Matched cohorts with prior infection*** |  |  |
| Total follow-up time (person-weeks) | 12,212 | 12,222 |
| Number of incident infections | 18 | 12 |
| Incidence rate of infection (per 10,000 person-weeks; 95% CI) | 14.7 (9.3 to 23.4) | 9.8 (5.6 to 17.3) |
| Unadjusted hazard ratio for infection with an omicron subvariant (95% CI) | 1.50 (0.72 to 3.11) |  |
| Adjusted hazard ratio for infection with an omicron subvariant (95% CI)<sup>†</sup> | 1.50 (0.72 to 3.12) |  |
| Effectiveness against infection with an omicron subvariant in % (95% CI)<sup>‡</sup> | -33.3 (-68.0 to 27.5) |  |
| ***Matched cohorts with prior pre-omicron infection*** |  |  |
| Total follow-up time (person-weeks) | 7,220 | 7,234 |
| Number of incident infections | 13 | 7 |
| Incidence rate of infection (per 10,000 person-weeks; 95% CI) | 18.0 (10.5 to 31.0) | 9.7 (4.6 to 20.3) |
| Unadjusted hazard ratio for infection with an omicron subvariant (95% CI) | 1.86 (0.74 to 4.65) |  |
| Adjusted hazard ratio for infection with an omicron subvariant (95% CI)<sup>†</sup> | 1.97 (0.78 to 4.95) |  |
| Effectiveness against infection with an omicron subvariant in % (95% CI)<sup>‡</sup> | -49.2 (-79.8 to 21.7) |  |
| ***Matched cohorts with prior omicron infection*** |  |  |
| Total follow-up time (person-weeks) | 4,901 | 4,897 |
| Number of incident infections | 5 | 5 |
| Incidence rate of infection (per 10,000 person-weeks; 95% CI) | 10.2 (4.3 to 24.5) | 10.2 (4.3 to 24.5) |
| Unadjusted hazard ratio for infection with an omicron subvariant (95% CI) | 1.00 (0.29 to 3.45) |  |
| Adjusted hazard ratio for infection with an omicron subvariant (95% CI)<sup>†</sup> | 0.99 (0.29 to 3.42) |  |
| Effectiveness against infection with an omicron subvariant in % (95% CI)<sup>‡</sup> | 1.2 (-70.7 to 71.4) |  |
| ***Full matched cohorts with and with no prior infection*** |  |  |
| Total follow-up time (person-weeks) | 72,442 | 71,826 |
| Number of incident infections | 127 | 152 |
| Incidence rate of infection (per 10,000 person-weeks; 95% CI) | 17.7 (14.9 to 21.0) | 21.0 (17.9 to 24.6) |
| Unadjusted hazard ratio for infection with an omicron subvariant (95% CI) | 0.83 (0.65 to 1.05) |  |
| Adjusted hazard ratio for infection with an omicron subvariant (95% CI)<sup>†</sup> | 0.82 (0.65 to 1.04) |  |
| Effectiveness against infection with an omicron subvariant in % (95% CI)<sup>‡</sup> | 17.7 (-4.1 to 35.0) |  |

CI denotes confidence interval.
\*Each adolescent vaccinated with the 30-µg BNT162b2 vaccine was matched exactly one-to-one by sex, 10 nationality groups, number of coexisting conditions, prior infection status, and calendar month of the second vaccine dose to the first eligible adolescent vaccinated with the pediatric 10-µg BNT162b2 vaccine who was alive and did not have a SARS-CoV-2 positive test in the 90 days before the start of the follow-up (14 days after the second vaccine dose of their match).
<sup>†</sup>Cox regression analysis adjusted for sex, 10 nationality groups, number of coexisting conditions, prior infection status, and calendar month of the second vaccine dose.

Cumulative incidence of infection was 6.0% (95% CI: 4.9-7.3%) for the 30-µg cohort and 7.2% (95% CI: 6.1-8.5%) for the 10-µg cohort, 210 days after the start of follow-up (Figure 1).

Incidence during follow-up was dominated by omicron subvariants including, consecutively, BA.1/BA.2,^28^ BA.4/BA.5,^29^ BA.2.75* (predominantly BA.2.75.2),^30^ and very recently XBB.

Adjusted hazard ratio comparing incidence of infection in the 30-µg cohort to the 10-µg cohort was 0.77 (95% CI: 0.60-0.98; Table 2). Corresponding relative effectiveness was 23.4% (95% CI: 1.6-40.4%).

The proportion of adolescents who had a SARS-CoV-2 test during follow-up was 66.8% for the 30-µg cohort and 70.7% for the 10-µg cohort. The testing frequency was 1.08 and 1.13 tests per person, respectively. Effectiveness additionally adjusted in a sensitivity analysis for the differences in the testing frequency between the cohorts was 19.5% (−3.3-37.4%).

### Additional relative effectiveness measures

Relative effectiveness was -33.3% (95% CI: -68.0-27.5%), -49.2% (95% CI: -79.8-21.7%), and 1.2% (95% CI: -70.7-71.4%) by restricting the cohorts to each of persons with any prior infection, only prior pre-omicron infection, and only prior omicron infection, respectively (Table 2). Relative effectiveness was 17.7% (95% CI: -4.1-35.0%) in the full matched cohorts including both persons with and with no prior infections (Table 2).

## Discussion

The 30-µg antigen dosage was associated with ∼25% reduction in incidence of omicron infection compared to the 10-µg antigen dosage among adolescents with no record of prior infection.

These findings may be explained by the larger dosage inducing a larger volume of antibodies.^31^ Strikingly, this effect size for the difference in protection is similar to that observed (∼30%) for the protection of the mRNA-1273 vaccine relative to the BNT162b2 vaccine,^3^ where the difference in antigen dosage is also about three-fold (100 µg^1^ versus 30 µg^2^, respectively).

The estimate for the increased protection induced by the 30-µg antigen dosage is perhaps conservative. There was a difference of 8.4 months in age between the cohorts, and since vaccine effectiveness appears stronger at younger age,^4^ possibly because of a smaller body weight, the effect size may have been slightly higher if the comparison was done in persons with the exact same age. Though those with a documented prior infection were excluded from the analysis, some of those with no record of prior infection may have been infected but their infections were never documented. If prior infection levels the differences in protection induced by vaccination, as seems suggested by the results of the additional analyses in this study and considering that natural immunity induces stronger and more durable protection against infection than vaccine immunity,^32^ the undocumented prior infections may have biased the estimate towards smaller differences in protection.

This study has limitations. The sizes of the subcohorts for those with a documented prior infection were much smaller than those with no documented prior infection leading to wide 95% CIs for the differences in incidence. With the lower severity of SARS-CoV-2 infection among adolescents^33,34^ and the lower severity of omicron infections,^35,36^ no infections progressed to severe,^12^ critical,^12^ and fatal^13^ COVID-19 in any of the cohorts to estimate differences in protection against severe forms of COVID-19.

We investigated incidence of documented infections, but other infections may have occurred and gone undocumented. Home-based rapid antigen testing is not documented, and thus not factored in the analysis. However, there is no reason to believe that home-based testing could have differentially affected the followed cohorts to affect our effectiveness estimates. Matching was done to control for confounders known to affect infection exposure in Qatar,^10,17-20^ and this may have also controlled or reduced any differences in home-based testing between cohorts, given that it factored the key socio-demographic factors of the population of Qatar.

As an observational study, investigated cohorts were neither blinded nor randomized, so unmeasured or uncontrolled confounding cannot be excluded. Although matching covered key factors affecting infection exposure,^10,17-20^ it was not possible for other factors such as geography, for which data were unavailable. However, Qatar is essentially a city state and infection incidence was broadly distributed across neighborhoods.

The matching procedure used in this study was investigated in previous studies of different epidemiologic designs, and using control groups to test for null effects.^3,9,21-23^ These control groups have included unvaccinated cohorts versus vaccinated cohorts within two weeks of the first dose^9,21-23^ (when vaccine protection is negligible),^1,2^ and mRNA-1273-versus BNT162b2-vaccinated cohorts, also in the first two weeks after the first dose.^3^ These prior studies demonstrated at different times during the pandemic that this procedure provides adequate control of differences in infection exposure,^3,9,21-23^ suggesting that the matching strategy may also have controlled for differences in infection exposure in these studies.

In conclusion, a three-fold higher BNT162b2 antigen dosage was associated with ∼25% higher protection against omicron infection in adolescents of similar age and with no record of prior infection. These findings may inform design of future COVID-19 vaccines and boosters for persons of different age groups.

## Data Availability

The dataset of this study is a property of the Qatar Ministry of Public Health that was provided to the researchers through a restricted-access agreement that prevents sharing the dataset with a third party or publicly. Future access to this dataset can be considered through a direct application for data access to Her Excellency the Minister of Public Health (https://www.moph.gov.qa/english/Pages/default.aspx). Aggregate data are available within the manuscript and its Supplementary information.

## Funding

We acknowledge the many dedicated individuals at Hamad Medical Corporation, the Ministry of Public Health, the Primary Health Care Corporation, Qatar Biobank, Sidra Medicine, and Weill Cornell Medicine-Qatar for their diligent efforts and contributions to make this study possible.

The authors are grateful for institutional salary support from the Biomedical Research Program and the Biostatistics, Epidemiology, and Biomathematics Research Core, both at Weill Cornell Medicine-Qatar, as well as for institutional salary support provided by the Ministry of Public Health, Hamad Medical Corporation, and Sidra Medicine. The authors are also grateful for the Qatar Genome Programme and Qatar University Biomedical Research Center for institutional support for the reagents needed for the viral genome sequencing. The funders of the study had no role in study design, data collection, data analysis, data interpretation, or writing of the article.

Statements made herein are solely the responsibility of the authors.

## Author contributions

HC co-designed the study, performed the statistical analyses, and co-wrote the first draft of the article. LJA conceived and co-designed the study, led the statistical analyses, and co-wrote the first draft of the article. PVC designed mass PCR testing to allow routine capture of SGTF variants. PT and MRH conducted the multiplex, RT-qPCR variant screening and viral genome sequencing. HY, HAK, and AAA conducted viral genome sequencing. All authors contributed to data collection and acquisition, database development, discussion and interpretation of the results, and to the writing of the manuscript. All authors have read and approved the final manuscript.

## Supplementary Appendix

## Section S1. Further details on methods

### Data sources and testing

Severe acute respiratory syndrome coronavirus 2 (SARS-CoV-2) testing in the healthcare system in Qatar is done at a mass scale, and mostly for routine reasons, where about 5% of the population were tested every week,^1,2^ up to October 31, 2022. About 75% of those diagnosed are diagnosed not because of appearance of symptoms, but because of routine testing.^1,2^ Every polymerase chain reaction (PCR) test and an increasing proportion of the facility-based rapid antigen tests conducted in Qatar, regardless of location or setting, are classified on the basis of symptoms and the reason for testing (clinical symptoms, contact tracing, surveys or random testing campaigns, individual requests, routine healthcare testing, pre-travel, at port of entry, or other). All facility-based testing done during follow-up in the present study was factored in the analyses of this study.

Rapid antigen test kits are available for purchase in pharmacies in Qatar, but outcome of home-based testing is not reported nor documented in the national databases. Since SARS-CoV-2-test outcomes are linked to specific public health measures, restrictions, and privileges, testing policy and guidelines stress facility-based testing as the core testing mechanism in the population.

While facility-based testing is provided free of charge or at low subsidized costs, depending on the reason for testing, home-based rapid antigen testing is de-emphasized and not supported as part of national policy. There is no reason to believe that home-based testing could have differentially affected the followed matched cohorts to affect our results.

Qatar has unusually young, diverse demographics, in that only 9% of its residents are ≥50 years of age, and 89% are expatriates from over 150 countries.^3,4^ Further descriptions of the study population and these national databases were reported previously.^1,2,4-6^

### Comorbidity classification

Comorbidities were ascertained and classified based on the ICD-10 codes for chronic conditions as recorded in the electronic health record encounters of each individual in the Cerner-system national database that includes all citizens and residents registered in the national and universal public healthcare system. All encounters for each individual were analyzed to determine the comorbidity classification for that individual, as part of a recent national analysis to assess healthcare needs and resource allocation. The Cerner-system national database includes encounters starting from 2013, after this system was launched in Qatar. As long as each individual had at least one encounter with a specific comorbidity diagnosis since 2013, this person was classified with this comorbidity. Individuals who have comorbidities but never sought care in the public healthcare system, or seek care exclusively in private healthcare facilities, are classified as individuals with no comorbidity due to absence of recorded encounters for them.

### Section S2. Laboratory methods and variant ascertainment

#### Real-time reverse-transcription polymerase chain reaction testing

Nasopharyngeal and/or oropharyngeal swabs were collected for polymerase chain reaction (PCR) testing and placed in Universal Transport Medium (UTM). Aliquots of UTM were: 1) extracted on KingFisher Flex (Thermo Fisher Scientific, USA), MGISP-960 (MGI, China), or ExiPrep 96 Lite (Bioneer, South Korea) followed by testing with real-time reverse-transcription PCR (RT-qPCR) using TaqPath COVID-19 Combo Kits (Thermo Fisher Scientific, USA) on an ABI 7500 FAST (Thermo Fisher Scientific, USA); 2) tested directly on the Cepheid GeneXpert system using the Xpert Xpress SARS-CoV-2 (Cepheid, USA); or 3) loaded directly into a Roche cobas 6800 system and assayed with the cobas SARS-CoV-2 Test (Roche, Switzerland). The first assay targets the viral S, N, and ORF1ab gene regions. The second targets the viral N and E-gene regions, and the third targets the ORF1ab and E-gene regions.

All PCR testing was conducted at the Hamad Medical Corporation Central Laboratory or Sidra Medicine Laboratory, following standardized protocols.

#### Rapid antigen testing

Severe acute respiratory syndrome coronavirus 2 (SARS-CoV-2) antigen tests were performed on nasopharyngeal swabs using one of the following lateral flow antigen tests: Panbio COVID-19 Ag Rapid Test Device (Abbott, USA); SARS-CoV-2 Rapid Antigen Test (Roche, Switzerland); Standard Q COVID-19 Antigen Test (SD Biosensor, Korea); or CareStart COVID-19 Antigen Test (Access Bio, USA). All antigen tests were performed point-of-care according to each manufacturer’s instructions at public or private hospitals and clinics throughout Qatar with prior authorization and training by the Ministry of Public Health (MOPH). Antigen test results were electronically reported to the MOPH in real time using the Antigen Test Management System which is integrated with the national Coronavirus Disease 2019 (COVID-19) database.

#### Classification of infections by variant type

Surveillance for the severe acute respiratory syndrome coronavirus 2 (SARS-CoV-2) variants in Qatar is based on viral genome sequencing and multiplex real-time reverse-transcription polymerase chain reaction (RT-qPCR) variant screening^7^ of random positive clinical samples,^2,8-12^ complemented by deep sequencing of wastewater samples.^10,13,14^ Further details on the viral genome sequencing and multiplex RT-qPCR variant screening throughout the SARS-CoV-2 waves in Qatar can be found in previous publications.^1,2,6,8-12,15-20^

### Section S3. COVID-19 severity, criticality, and fatality classification

Classification of Coronavirus Disease 2019 (COVID-19) case severity (acute-care hospitalizations),^21^ criticality (intensive-care-unit hospitalizations),^21^ and fatality^22^ followed World Health Organization (WHO) guidelines. Assessments were made by trained medical personnel independent of study investigators and using individual chart reviews, as part of a national protocol applied to every hospitalized COVID-19 patient. Each hospitalized COVID-19 patient underwent an infection severity assessment every three days until discharge or death. We classified individuals who progressed to severe, critical, or fatal COVID-19 between the time of the documented infection and the end of the study based on their worst outcome, starting with death,^22^ followed by critical disease,^21^ and then severe disease.^21^

Severe COVID-19 disease was defined per WHO classification as a SARS-CoV-2 infected person with “oxygen saturation of <90% on room air, and/or respiratory rate of >30 breaths/minute in adults and children >5 years old (or ≥60 breaths/minute in children <2 months old or ≥50 breaths/minute in children 2-11 months old or ≥40 breaths/minute in children 1–5 years old), and/or signs of severe respiratory distress (accessory muscle use and inability to complete full sentences, and, in children, very severe chest wall indrawing, grunting, central cyanosis, or presence of any other general danger signs)”.^21^ Detailed WHO criteria for classifying Severe acute respiratory syndrome coronavirus 2 (SARS-CoV-2) infection severity can be found in the WHO technical report.^21^

Critical COVID-19 disease was defined per WHO classification as a SARS-CoV-2 infected person with “acute respiratory distress syndrome, sepsis, septic shock, or other conditions that would normally require the provision of life sustaining therapies such as mechanical ventilation (invasive or non-invasive) or vasopressor therapy”.^21^ Detailed WHO criteria for classifying SARS-CoV-2 infection criticality can be found in the WHO technical report.^21^

COVID-19 death was defined per WHO classification as “a death resulting from a clinically compatible illness, in a probable or confirmed COVID-19 case, unless there is a clear alternative cause of death that cannot be related to COVID-19 disease (e.g. trauma). There should be no period of complete recovery from COVID-19 between illness and death. A death due to COVID-19 may not be attributed to another disease (e.g. cancer) and should be counted independently of preexisting conditions that are suspected of triggering a severe course of COVID-19”. Detailed WHO criteria for classifying COVID-19 death can be found in the WHO technical report.^22^

**Figure S1.**
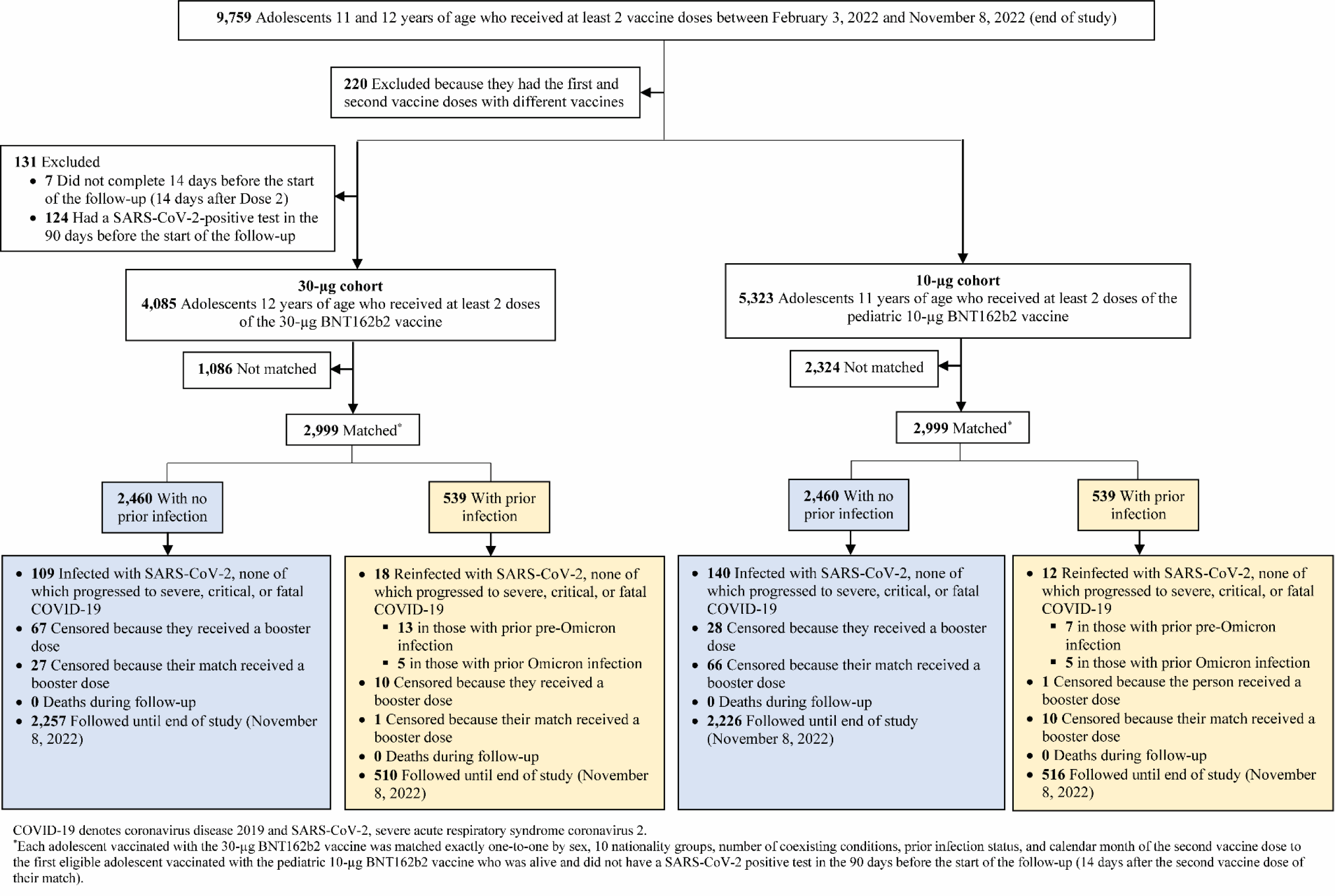
Cohort selection for investigating relative effectiveness of the 30-µg BNT162b2 vaccine compared to the pediatric 10-µg BNT162b2 vaccine against infection with an omicron subvariant.

**Table S1.** STROBE checklist for cohort studies.

|  | Item No | Recommendation | Main Text page |
| --- | --- | --- | --- |
| Title and abstract | 1 | (a) Indicate the study’s design with a commonly used term in the title or the abstract | Abstract |
|  |  | (b) Provide in the abstract an informative and balanced summary of what was done and what was found | Abstract |
| Introduction |  |  |  |
| Background/rationale | 2 | Explain the scientific background and rationale for the investigation being reported | Introduction |
| Objectives | 3 | State specific objectives, including any prespecified hypotheses | Introduction |
| Methods |  |  |  |
| Study design | 4 | Present key elements of study design early in the paper | Methods (‘Study design and cohorts’) |
| Setting | 5 | Describe the setting, locations, and relevant dates, including periods of recruitment, exposure, follow-up, and data collection | Methods (‘Study design and cohorts’) & Figure S1 in Supplementary Appendix |
| Participants | 6 | (a) Give the eligibility criteria, and the sources and methods of selection of participants. Describe methods of follow-up<br>(b) For matched studies, give matching criteria and number of exposed and unexposed | Methods (‘Study design and cohorts’ & ‘Cohort matching and follow-up’) & Figure S1 in Supplementary Appendix |
| Variables | 7 | Clearly define all outcomes, exposures, predictors, potential confounders, and effect modifiers. Give diagnostic criteria, if applicable | Methods (‘Study design and cohorts’ & ‘Cohort matching and follow-up’), Table 1, & Sections S1-S3 in Supplementary Appendix |
| Data sources/measurement | 8* | For each variable of interest, give sources of data and details of methods of assessment (measurement). Describe comparability of assessment methods if there is more than one group | Methods (‘Study population and data sources’ & ‘Statistical analysis’, paragraph 1), Table 1, & Sections S1-S3 in Supplementary Appendix |
| Bias | 9 | Describe any efforts to address potential sources of bias | Methods (‘Cohort matching and follow-up’ & ‘Statistical analysis’) |
| Study size | 10 | Explain how the study size was arrived at | Figure S1 |
| Quantitative variables | 11 | Explain how quantitative variables were handled in the analyses. If applicable, describe which groupings were chosen and why | Methods (‘Cohort matching and follow-up’) & Table 1 |
| Statistical methods | 12 | (a) Describe all statistical methods, including those used to control for confounding | Methods (‘Statistical analysis’) |
|  |  | (b) Describe any methods used to examine subgroups and interactions | Methods (‘Statistical analysis’, paragraph 5) |
|  |  | (c) Explain how missing data were addressed | Not applicable, see Methods (‘Study population and data sources’) |
|  |  | (d) If applicable, explain how loss to follow-up was addressed | Not applicable, see Methods (‘Study population and data sources’) |
|  |  | (e) Describe any sensitivity analyses | Methods (‘Statistical analysis’, paragraph 2) |
| Results |  |  |  |
| Participants | 13* | (a) Report numbers of individuals at each stage of study—eg numbers potentially eligible, examined for eligibility, confirmed eligible, included in the study, completing follow-up, and analysed<br>(b) Give reasons for non-participation at each stage<br>(c) Consider use of a flow diagram | Results (‘Study population’), Figure S1, & Table 1 |
| Descriptive data | 14 | (a) Give characteristics of study participants (eg demographic, clinical, social) and information on exposures and potential confounders | Results (‘Study population’), Table 1, & Figure S1 in Supplementary Appendix |
|  |  | (b) Indicate number of participants with missing data for each variable of interest | Not applicable, see Methods (‘Study population and data sources’) |
|  |  | (c) Summarise follow-up time (eg, average and total amount) | Results (‘Relative effectiveness in adolescents with no prior infection’, paragraph 3), Figure 1, & Table 2 |
| Outcome data | 15 | Report numbers of outcome events or summary measures over time | Results (‘Relative effectiveness in adolescents with no prior infection', paragraphs 3-4), Figure 1, Table 2, & Table S1 in Supplementary appendix. |
| Main results | 16 | (a) Give unadjusted estimates and, if applicable, confounder-adjusted estimates and their precision (eg, 95% confidence interval). Make clear which confounders were adjusted for and why they were included | Results ('Relative effectiveness in adolescents with no prior infection', paragraph 5) & Table 2 |
|  |  | (b) Report category boundaries when continuous variables were categorized | Table 1 |
|  |  | (c) If relevant, consider translating estimates of relative risk into absolute risk for a meaningful time period | Not applicable |
| Other analyses | 17 | Report other analyses done—eg analyses of subgroups and interactions, and sensitivity analyses | Results ('Additional relative effectiveness measures') & Table 2 |
| Discussion |  |  |  |
| Key results | 18 | Summarise key results with reference to study objectives | Discussion, paragraphs 1-2 |
| Limitations | 19 | Discuss limitations of the study, taking into account sources of potential bias or imprecision. Discuss both direction and magnitude of any potential bias | Discussion, paragraphs 3-6 |
| Interpretation | 20 | Give a cautious overall interpretation of results considering objectives, limitations, multiplicity of analyses, results from similar studies, and other relevant evidence | Discussion, paragraph 7 |
| Generalisability | 21 | Discuss the generalisability (external validity) of the study results | Discussion, paragraphs 3-6 |
| Other information |  |  |  |
| Funding | 22 | Give the source of funding and the role of the funders for the present study and, if applicable, for the original study on which the present article is based | Funding |

## Notes

### Competing Interest Statement

The authors have declared no competing interest.

### Author Declarations

The institutional review boards at Hamad Medical Corporation and Weill Cornell Medicine in Qatar approved this retrospective study with a waiver of informed consent.

